# Proteome-wide Mendelian randomization identifies causal links between blood proteins and severe COVID-19

**DOI:** 10.1101/2021.03.09.21253206

**Authors:** Alish B. Palmos, Vincent Millischer, David K. Menon, Timothy R. Nicholson, Leonie Taams, Benedict Michael, COVID Clinical Neuroscience Study Consortium, Christopher Hübel, Gerome Breen

**Affiliations:** Social, Genetic & Developmental Psychiatry Centre, Institute of Psychiatry, Psychology & Neuroscience, King’s College London, UK; UK National Institute for Health Research (NIHR) Biomedical Research Centre for Mental Health, South London and Maudsley Hospital, London, UK; Department of Psychiatry and Psychotherapy, Medical University of Vienna, Vienna, Austria; Department of Molecular Medicine and Surgery, Karolinska Institutet, Stockholm, Sweden; Division of Anaesthesia, Department of Medicine, University of Cambridge; Centre for Inflammation Biology & Cancer Immunology, Department of Inflammation Biology, School of Immunology & Microbial Sciences, King’s College London, UK; Institute of Infection and Global Health, University of Liverpool, UK; National Centre for Register-based Research, Department of Economics and Business Economics, Aarhus University, Aarhus, Denmark; Institute of Psychiatry, Psychology & Neuroscience, King’s College London, UK

## Abstract

The COVID-19 pandemic death toll now surpasses two million individuals and there is a need for early identification of individuals at increased risk of mortality. Host genetic variation partially drives the immune and biochemical responses to COVID-19 that lead to risk of mortality. We identify and prioritise blood proteins and biomarkers that may indicate increased risk for severe COVID-19, via a proteome Mendelian randomization approach by collecting genome-wide association study (GWAS) summary statistics for >4,000 blood proteins. After multiple testing correction, troponin I3, cardiac type (TNNI3) had the strongest effect (odds ratio (O.R.) of 6.86 per standard deviation increase in protein level), with proteinase 3 (PRTN3) (O.R.=2.48), major histocompatibility complex, class II, DQ alpha 2 (HLA-DQA2) (O.R.=2.29), the C4A-C4B heterodimer (O.R.=1.76) and low-density lipoprotein receptor-related protein associated protein 1 (LRPAP1) (O.R.=1.73) also being associated with higher odds of severe COVID-19. Conversely, major histocompatibility complex class I polypeptide-related sequence A (MHC1A) (O.R.=0.6) and natural cytotoxicity triggering receptor 3 (NCR3) (O.R.=0.46) were associated with lower odds. These proteins are involved in heart muscle contraction, natural killer and antigen presenting cells, and the major histocompatibility complex. Based on these findings, it may be possible to better predict which patients may develop severe COVID-19 and to design better treatments targeting the implicated mechanisms.

## Introduction

The severe acute respiratory syndrome coronavirus 2 (SARS-CoV-2) was identified in late 2019 in Wuhan, China, as the cause of what is now termed the coronavirus disease 2019 (COVID-19) that rapidly evolved into a global pandemic (1,2). As of February 2021, more than 100 million cases have been confirmed worldwide with total deaths exceeding 2 million (3). COVID-19 pathology encompasses a wide spectrum of clinical manifestations from asymptomatic, over mild, moderate, to severe infections (around 15%) (1,2). Severe COVID-19 commonly requires hospitalization and intensive care with assisted respiratory support, and subsequent respiratory failure is the most common reason for COVID-19 associated mortality (4).

A dysregulated pro- and anti-inflammatory immunomodulatory response drives much of the severe pathophysiology of COVID-19 and comprises alveolar damage, lung inflammation and pathology of an acute respiratory distress syndrome (1,2)^,(5,6)^. While initial descriptions and a response to corticosteroids led to a label of a “cytokine storm” (7) in severe disease cases, recent reports suggest that the abnormalities in host response may be more complex and nuanced (8). Given that the innate immune response has an individual-level genetic basis, genetic variants carried by an individual could play an important role in the individual-level immune response and, therefore, may influence progression and severity of COVID-19. Most biological systems are based on the interaction of biological and environmental factors. The influence of environmental factors can confound measured associations between biological markers and diseases outcomes. Therefore, studying immunomodulatory blood proteins in COVID-19 patients is difficult due to potential confounding effects of factors, such as initial viral exposure/inoculum, smoking behaviour and high body mass index (BMI). These factors themselves are associated with increased pro-inflammatory cytokine levels and may represent independent risk factors for severe COVID-19 (9–11).

Numerous genome-wide association studies (GWASs) in healthy populations associated genetic variants with immunomodulatory blood proteins (12–14). In addition, the COVID-19 Host Genetics Initiative carried out GWASs on COVID-19 subtypes to understand the role of host genetic factors in susceptibility and severity of COVID-19 (15). These data represent a powerful source of information to find new biomarkers and therapeutic leads. The method of Mendelian randomization can be applied to investigate the relationship between immunomodulatory blood proteins and severe COVID-19 infection. This exploits the fact that alleles are randomly inherited from parent to offspring in a manner analogous to a randomised-controlled trial, and allows estimation of putative causal effects of an exposure on a disease while avoiding confounding environmental effects, thus overcoming some of the limitations of observational studies. Recent advancements in Mendelian randomization methods allow use of GWAS summary statistics data, to identify genetic proxies (i.e., instrumental variables) of modifiable risk factors and testing their association with disease outcomes (16). We, therefore, conducted Mendelian randomization analyses between the levels of a large number of blood proteins and severe COVID-19 to identify putative causal associations that could be prioritised in clinical studies.

## Methods

### Blood protein GWAS data

In total, we amassed 5,305 sets of GWAS summary statistics for blood biomarkers (12,13,84–89). A systematic search was performed based on the ontology lookup service (OLS; www.ebi.ac.uk/ols/index) using R and the packages ‘rols’ and ‘gwasrapid’ between July 7th and July 27th, 2020. OLS is a repository for biomedical ontologies, such as gene ontology (GO) or the experimental factor ontology (EFO), including a systematic description of many experimental variables. First, all subnodes of the EFO ‘protein measurements’ (EFO:0004747) were determined using an iterative process based on the package ‘rols’. Overall, 628 unique EFO IDs were determined. Subsequently, all genetic associations reported in the GWAS Catalog (90) (www.ebi.ac.uk/gwas/) were determined and linked to the corresponding study using the package ‘gwasrapid’. One hundred and seventy-eight unique GWAS catalog accession IDs with available summary statistics were curated manually before inclusion. In order to expand the dataset, studies published at a later date were included manually and the first and the last author of studies without publicly available summary statistics were contacted.

In total, we included eight publications for which summary data was readily available and processed those using standard GWAS summary statistics quality control metrics including removal of incomplete genetic variants, variants with information metrics of lower than 0.6 and allele frequencies more extreme than 0.005 or 0.995. Allele frequencies were taken from the 1,000 Genomes Project data where needed (91). See Supplementary Table 1 for a full list of the studies included in these analyses.

### COVID-19 GWAS data

We downloaded the COVID-19 Host Genome Initiative GWAS meta-analysis (15) of “very severe respiratory confirmed covid vs not hospitalized covid” (release 4, October 2020; www.covid19hg.org/results/). Cases and controls were defined as SARS-CoV-2 infected individuals within a clinical window of 15-90 days from diagnosis; with cases defined as mortality or the need for respiratory support (intubation, continuous positive airway pressure or bilevel positive airway pressure) and controls defined as surviving individuals who did not require respiratory support at any point since diagnosis.(15) The European sample consisted of 269 cases and 688 controls.

This GWAS was selected due to its extreme phenotyping, focusing specifically on mortality and respiratory failure (cases) compared to discharge from hospital without respiratory complications (controls). Subsequent COVID-19 GWAS are based on different, much less severe outcomes and broader phenotypes such as hospitalisation versus non hospitalisation that do not directly address the encompassing genetic basis for COVID-19 mortality or respiratory failure.

### Mendelian randomization

To examine the influence of blood proteins on the risk of developing severe COVID-19, we selected genetic variants, predominantly single nucleotide polymorphisms (SNPs), that were strongly associated with actual blood protein levels in 3,609 genome-wide analyses of single proteins using robust methodologies (see supplement for more details on how the proteins were measured). Using these genetic loci as proxies for protein levels we performed an analysis using Mendelian randomization, a method that enables tests of putative causal associations of these blood proteins with the development of severe COVID-19. We used the Generalised Summary-based Mendelian randomization (GSMR) method as the base method (92).

For all GWASs, SNPs were used as instrumental variables and were selected by applying a suggestive *p-value* threshold (*p* < 5 × 10^−6^), in order to identify enough SNPs in common between the exposure (i.e., blood marker) and outcome (i.e. severe COVID-19). Where possible (due to SNPs in common between the exposure and the outcome), bidirectional analyses were performed between 3,609 blood markers and severe COVID-19, resulting in a total of 7,406 tests. To account for multiple testing, we calculated false discovery rate (FDR) corrected *Q* values using the p.adjust function in R (*p*_FDR_ = 0.1).(93)

### Sensitivity analyses

To test for robustness, we performed sensitivity analyses on our significant results from the GSMR analyses using additional Mendelian randomization methods, including the weighted median (WM), inverse variance-weighted (IVW), robust adjusted profile score (MR-RAPS), and MR-Egger methods (94–96). In order to pass our sensitivity analyses, at least three of these four methods must agree with the GSMR results.

### Clinical application

The protein with the highest effect size was investigated for its clinical application. For this we used the Open Targets Platform (20). Specifically we focus on gene ontology and drug targets.

## Results

Out of the possible 7,406 associations tested, we tested 3,609 associations with severe COVID-19 as the exposure and 3,797 associations with severe COVID-19 as the outcome. No tests with severe COVID-19 as the exposure reached a level of statistical significance and are therefore not reported. See Table 1 for significantly associated blood markers alongside test statistics.

**Table 1.**
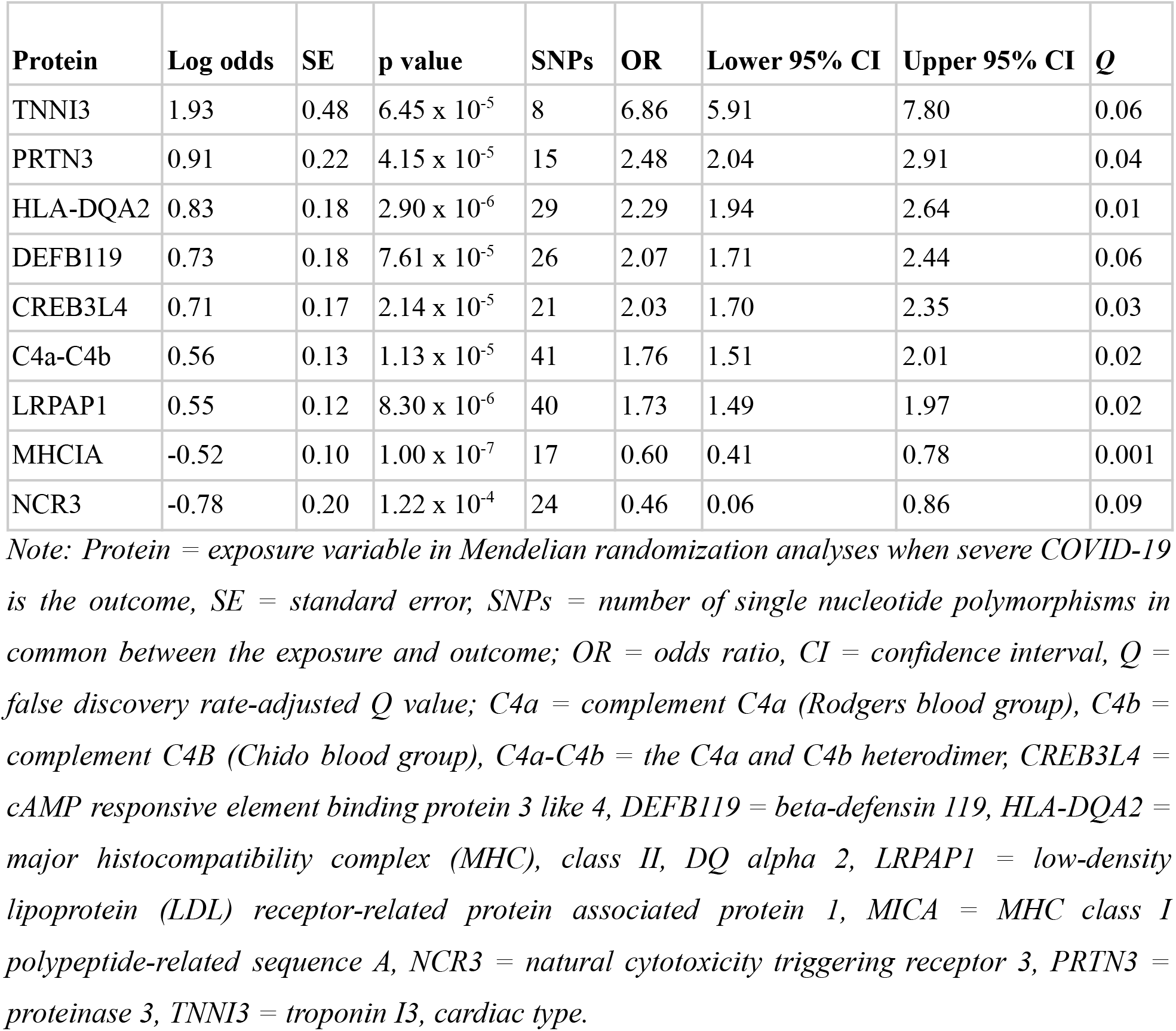
Mendelian randomization results. This table details significant, false discovery rate-(FDR)-corrected Mendelian randomization results, using the Generalised Summary Data-based Mendelian randomization (GSMR) method. Seven blood markers were associated with a significantly increased risk for severe COVID-19 while two blood markers were associated with a significantly decreased risk for severe COVID-19. CREB3L4 and DEFB119 did not survive our sensitivity analyses. The genetic instruments for C4a-C4b and HLA-DQA2 were in low to moderate linkage disequilibrium but this did not influence results (see Supplementary Table 4). The table presents the log odds statistics and corresponding standard error as well as odds ratios, 95% confidence intervals, and the FDR-adjusted Q values (p_FDR_ = 0.1).

| **Protein** | **Log odds** | **SE** | **p value** | **SNPs** | **OR** | **Lower 95% CI** | **Upper 95% CI** | **$Q$** |
| --- | --- | --- | --- | --- | --- | --- | --- | --- |
| TNNI3 | 1.93 | 0.48 | $6.45 \times 10^{-5}$ | 8 | 6.86 | 5.91 | 7.80 | 0.06 |
| PRTN3 | 0.91 | 0.22 | $4.15 \times 10^{-5}$ | 15 | 2.48 | 2.04 | 2.91 | 0.04 |
| HLA-DQA2 | 0.83 | 0.18 | $2.90 \times 10^{-6}$ | 29 | 2.29 | 1.94 | 2.64 | 0.01 |
| DEFB119 | 0.73 | 0.18 | $7.61 \times 10^{-5}$ | 26 | 2.07 | 1.71 | 2.44 | 0.06 |
| CREB3L4 | 0.71 | 0.17 | $2.14 \times 10^{-5}$ | 21 | 2.03 | 1.70 | 2.35 | 0.03 |
| C4a-C4b | 0.56 | 0.13 | $1.13 \times 10^{-5}$ | 41 | 1.76 | 1.51 | 2.01 | 0.02 |
| LRPAP1 | 0.55 | 0.12 | $8.30 \times 10^{-6}$ | 40 | 1.73 | 1.49 | 1.97 | 0.02 |
| MHCIA | -0.52 | 0.10 | $1.00 \times 10^{-7}$ | 17 | 0.60 | 0.41 | 0.78 | 0.001 |
| NCR3 | -0.78 | 0.20 | $1.22 \times 10^{-4}$ | 24 | 0.46 | 0.06 | 0.86 | 0.09 |

*Note: Protein = exposure variable in Mendelian randomization analyses when severe COVID-19 is the outcome, SE = standard error, SNPs = number of single nucleotide polymorphisms in common between the exposure and outcome; OR = odds ratio, CI = confidence interval, $Q$ = false discovery rate-adjusted $Q$ value; C4a = complement C4a (Rodgers blood group), C4b = complement C4B (Chido blood group), C4a-C4b = the C4a and C4b heterodimer, CREB3L4 = cAMP responsive element binding protein 3 like 4, DEFB119 = beta-defensin 119, HLA-DQA2 = major histocompatibility complex (MHC), class II, DQ alpha 2, LRPAP1 = low-density lipoprotein (LDL) receptor-related protein associated protein 1, MICA = MHC class I polypeptide-related sequence A, NCR3 = natural cytotoxicity triggering receptor 3, PRTN3 = proteinase 3, TNNI3 = troponin I3, cardiac type.*

### Elevated risk for severe COVID-19

After multiple testing correction (*p*_FDR_ = 0.1), seven blood markers were associated with a statistically significantly elevated risk of severe COVID-19 (see Figure 1). One standard deviation (SD) increase in troponin I, cardiac muscle (TNNI3) was associated with 589% (95% CI: 5.91-7.80, *q* = 0.06) higher odds of severe COVID-19. One SD increase in myeloblastin (PRTN3) was associated with 148% (95% CI: 2.04-2.91, *q* = 0.04) higher odds of severe COVID-19. One SD increase in major histocompatibility complex (MHC), class II, DQ alpha 2 (HLA-DQA2) was associated with 129% (95% CI: 1.94-2.64, *q* = 0.01) higher odds of severe COVID-19. One SD increase in complement components 4A and 4B (C4a-C4b) was associated with 76% (95% CI: 1.51-2.01, *q* = 0.02) higher odds of severe COVID-19. One SD increase in low-density lipoprotein (LDL) receptor-related protein associated protein 1 (LRPAP1) was associated with 73% (95% CI: 1.49-1.97, *q* = 0.02) higher odds of severe COVID-19 infection.

**Figure 1.**
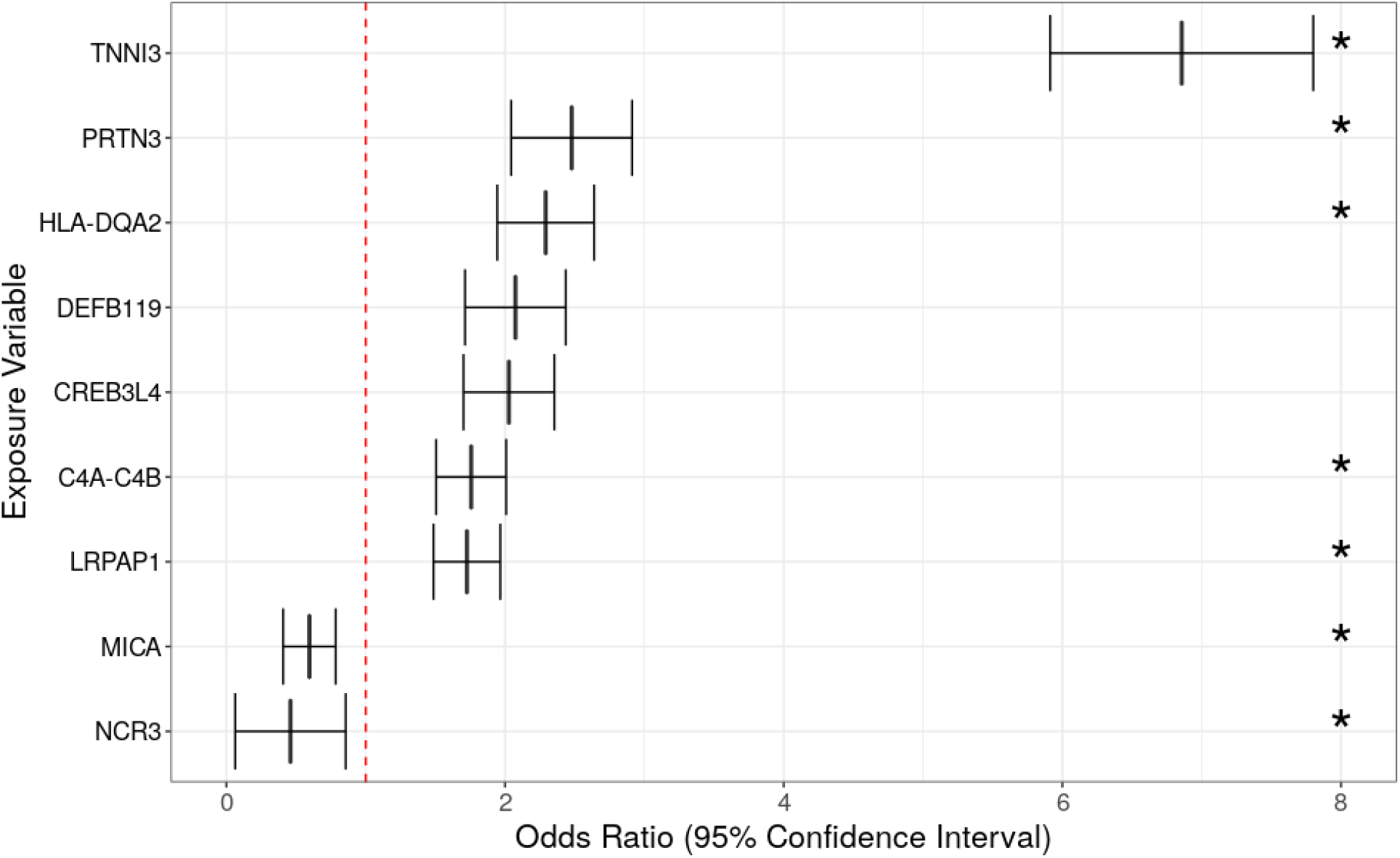
Blood markers putatively causally associated with severe COVID-19. Summary figure of the false discovery rate-corrected (p_FDR_ = 0.1) Mendelian randomization results using the Generalised Summary data-based Mendelian randomization (GSMR) method. Odds ratios (ORs) of the blood markers associated with severe COVID-19 are displayed on the x-axis (with 95% confidence intervals). The blood markers are displayed on the y-axis. The dashed red line represents an odds ratio of one (i.e., no effect). Seven blood markers were associated with a significantly increased risk for severe COVID-19 and two blood markers were associated with a significantly decreased risk for severe COVID-19 (q_FDR_ ≤ 0.1). Markers which additionally survived our sensitivity analyses are marked with an asterisk (*). C4A = complement C4A (Rodgers blood group), C4B = complement C4B (Chido blood group), CREB3L4 = cAMP responsive element binding protein 3 like 4, DEFB119 = beta-defensin 119, HLA-DQA2 = major histocompatibility complex (MHC), class II, DQ alpha 2, LRPAP1 = low-density lipoprotein (LDL) receptor-related protein associated protein 1, MICA = MHC class I polypeptide-related sequence A, NCR3 = natural cytotoxicity triggering receptor 3, PRTN3 = proteinase 3, TNNI3 = troponin I3, cardiac type.

### Decreased risk for severe COVID-19

A one SD increase in MHC class I polypeptide-related sequence A (MICA) was associated with 40% (95% CI: 0.41-0.78, *q* = 0.001) lower odds and one SD increase in natural cytotoxicity triggering receptor 3 (NCR3) was associated with 54% (95% CI: 0.06-0.86, *q* = 0.09) lower odds of severe COVID-19 (see Figure 1).

### Sensitivity analyses

To further increase confidence in the findings, we additionally filtered our results for those concurrent with results calculated by additional Mendelian randomization methods (WM, IVW and MR-RAPS). Our sensitivity analyses revealed that the TNNI3, PRTN3, HLA-DQA2, C4A-C4B, LRPAP1, MICA and NCR3 associations with severe COVID-19 were confirmed. The two additional markers, cAMP responsive element binding protein 3 like 4 (CREB3L4) and beta-defensin 119 (DEFB119), associated with an increased risk for severe COVID-19 with the GSMR method, did not survive our sensitivity analyses. See Figure 2 for Mendelian randomization scatter plots of the sensitivity analyses and Supplementary Table 2 for a full results table of the sensitivity analyses.

**Figure 2.**
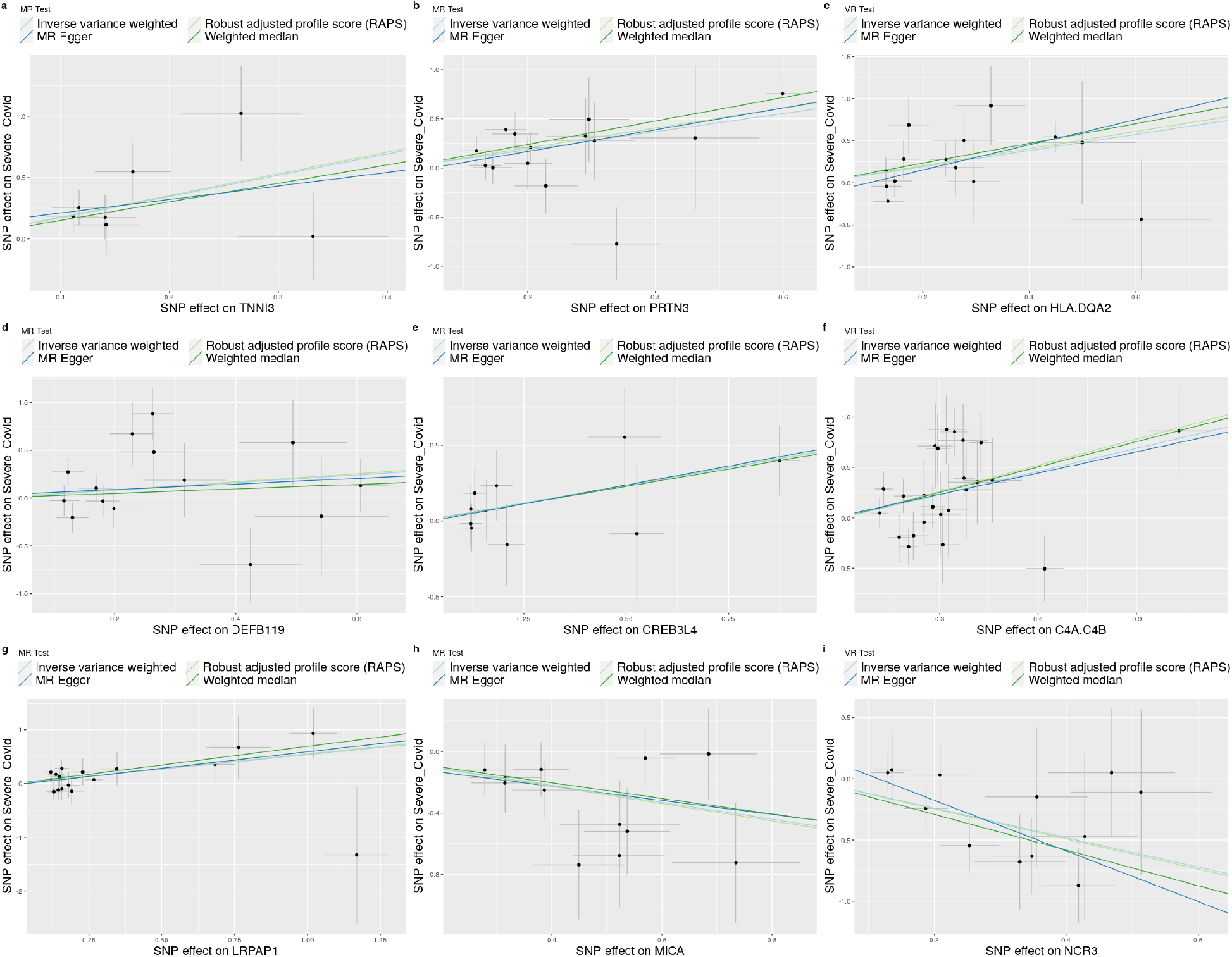
Sensitivity analyses for blood markers putatively causally associated with severe COVID-19. Mendelian randomization scatter plots using the inverse variance weighted, robust adjusted profile score, MR-Egger and weighted median methods associating each blood marker with severe COVID-19. Single nucleotide polymorphism (SNP) effects of the blood markers (i.e., exposures) are displayed on the x-axis and SNP effects of severe COVID-19 (outcome) are displayed on the y-axis. The horizontal line of each point represents the standard error of the SNP-exposure association and the vertical line the standard error of the SNP-outcome association. The effect of (A) TNNI3; (B) PRTN3; (C) HLA-DQ2; (D) DEFB119; (E) CREB3L4; (F) C4a-C4b; (G) LRPAP1; (H) MICA; (I) NCR3. SNP = single nucleotide polymorphism; C4A = complement C4A (Rodgers blood group), C4B = complement C4B (Chido blood group), CREB3L4 = cAMP responsive element binding protein 3 like 4, DEFB119 = beta-defensin 119, HLA-DQA2 = major histocompatibility complex (MHC), class II, DQ alpha 2, LRPAP1 = low-density lipoprotein (LDL) receptor-related protein associated protein 1, MICA = MHC class I polypeptide-related sequence A, NCR3 = natural cytotoxicity triggering receptor 3, PRTN3 = proteinase 3, TNNI3 = troponin I3, cardiac type.

Several of the proteins (MHC1A, NC3R, C4a-C4b, and HLA-DQA2) are encoded by non-overlapping genes in separate locations within the MHC (major histocompatibility complex) region, located on chromosome 6 (17–19). We verified that the associations observed are independent and not driven by linkage disequilibrium between the genetic instrumental variables used (see Supplementary Table 3). HLA proteins are vital in evoking immune responses to antigenic stimuli thereby orchestrating cellular and humoral immunity. These molecules facilitate the recognition of self or altered-self vs. non-self proteins by the body’s immune system (17–19).

### Clinical application

Due to its large effect, TNNI3 was further investigated for potential clinical application using the Open Targets Platform (20). We highlight the upstream effect that TNNI3 may be having on heart muscle contraction, which may be having a negative downstream effect on a wide range of biological processes in the nervous, respiratory and digestive systems (see Supplementary Figure 1 for a bubble plot of genetically associated processes with TNNI3). In addition, we identify levosimendan, a positive inotropic agent having ATP-dependent potassium-channel-opening and calcium-sensitizing effects by binding to a calcium sensing myocardial complex consisting of cardiac troponin C and troponin I(20)^,(21,22)^. This is a clinically approved drug used for heart failure, kidney failure and SARS (https://www.targetvalidation.org/summary?drug=CHEMBL2051955). We highlight the potential role levosimendan for severe COVID-19 (see Supplementary Figure 2 for a bubble plot of druggable targets associated with TNNI3 and levosimendan), but emphasise the need for further exploration of this relationship.

## Discussion

Our proteome-wide analysis of the influence of the blood levels of 3,609 proteins on risk for severe COVID-19 using Mendelian randomization, yielded evidence consistent with higher blood levels of TNNI3, PRTN3, HLADQA2, C4a-C4b and LRPAP1 being putative causal risk factors for severe COVID-19, and higher blood levels of MHC1A and NCR3 being protective against severe COVID-19. All of these proteins have detectable blood plasma or serum levels; we summarised functions of these blood markers in Supplementary Table 4. It is important to note that we did not identify the typical canonical immune proteins (such as interleukin-6 or C-reactive protein)(23,24), suggesting that with a larger database we may be able to pinpoint proteins that are upstream of major inflammatory mechanisms.

TNNI3, which codes for cardiac troponin I, showed the largest effect on COVID-19 severity in our Mendelian randomization analysis (OR per SD increase in TNNI3 level = 6.86, 95% C.I. 5.91-7.80). Cardiac troponin I is highly specific for heart muscle and higher concentrations in the blood are associated with higher general in-hospital mortality for a number of illnesses (25). It serves as a sensor of intracellular calcium, regulating the interaction between thick and thin sarcomere filaments during the heart muscle contraction (26). We hypothesise that this protein may be relevant to COVID-19 as ∼40% of deaths in a cohort from Wuhan (27) were attributed to myocardial damage and heart failure, as well as increased mortality in individuals with both COVID-19 and acute cardiac injury (28). Between 12-28% of patients with COVID-19 present with elevated troponin (29–32) are more likely to require intensive care (29,33), have higher in-hospital mortality(28,30,32,34–36) and show myocardial injury after recovery (37). Different cardiovascular complications, including myocarditis,(38) microangiopathy, myocardial infarction, or a dysregulated host immune response have been proposed to explain the high mortality in COVID-19(39) and troponin may be a predictor for mortality independent of inflammation.(40) In addition, TNNI3 has been associated with angiotensin-converting enzyme 2 (ACE2) upregulation in obstructive hypertrophic cardiomyopathy(41) and is shown to be a related target of ACE2(20). Given the importance of ACE2 in COVID-19,(42) this further implicates TNNI3 as an important target of severe disease pathophysiology (see Supplementary Table 5 for TNNI3 associated proteins). Levosimendan has been shown to exert action on the myocardial troponin complex,(20) (21,22)which involves complex interactions between troponin I and troponin C (43,44). Levosimendan is a calcium sensitiser, which canonically also exerts action on troponin C that is implicated in several diseases, including hypertrophic cardiomyopathy (45). Both troponin C and I (as well troponin T) are part of the troponin complex, with troponin I interacting with troponin C to regulate calcium sensitivity and contractility.(45) The translation of these findings to physiology is not straightforward since the effects of these target TNNI3 polymorphisms on the contribution of troponin I to myocardial Ca sensing are not established. Consequently, this finding may indicate either impaired calcium sensing in the presence of these SNPs, in which case a calcium sensitiser (such as levosimendan) may be helpful. Alternatively, these SNPs may drive greater Ca myocardial Ca sensitivity, and the increased blood levels of troponin I and poor outcomes seen in their presence may be due to myocardial necrosis in the presence of excessive myocardial stress – in which case Ca channel blockers might be indicated. In any event, our findings make a strong case for further exploration of this physiology and, potentially, clinical trials of promising interventions.(46)

MHC1A, also known as MICA, encodes a 62 kDa cell surface glycoprotein which is expressed on endothelial, dendritic, epithelial cells and fibroblasts. It is a target for cellular and humoral immune responses and becomes upregulated in response to stimuli of cellular stress like DNA damage and viral infection.(47) MHC1A binds to the natural killer group 2D (NKG2D) transmembrane protein activating natural killer cells, γδ T cells and CD8^+^ αβ T cells implicated in the antiviral response (48). Severe COVID-19 patients show high counts of NKG2C cells and low counts of NKG2D cells (49,50). Our findings suggest that a genetic propensity for higher blood MHC1A may protect against severe COVID-19, potentially through activating cytolytic cells (OR = 0.60, 95% CI: 0.41-0.78).

The natural cytotoxicity triggering receptor 3 (NCR3, also known as NKp30) is expressed by NK cells as well as by some peripheral blood CD8+ and γδ T cells and mediates NK cytotoxicity by facilitating the crosstalk between NK and dendritic cells.(51) NCR3 becomes upregulated in response to interferon γ and bacterial infection, suggesting a role in inflammatory and infectious diseases.(52) A separate transcriptomic analysis showed that reduced NCR3 transcription is associated with more severe influenza(53) while NCR3 plays important roles in both immunity against viral infections and in the development of immune evasion by viruses.(54) Our findings suggest that a genetic predisposition for higher blood NCR3 may protect against severe COVID-19 (OR = 0.46, 95% CI: 0.06-0.86).(55–57) However, we note that this association is with the soluble form of NCR3 found in plasma, complicating the interpretation of the results.

A one SD increase in levels of C4a-C4b was associated with COVID-19 severity (OR = 1.76, 95% CI 1.51-2.01). The C4a-C4b heterodimer is part of the complement cascade, which plays an important role in the host defense response against viral infections but has also been robustly associated with schizophrenia(58,59), with the associated C4 structural variants leading to possible altered neural C4A expression, excessive or inappropriate synaptic pruning(60,61). However, while a diagnosis of schizophrenia has been reported to be associated with a >7-fold increased risk for infection and almost a 3-fold increase in risk of mortality for COVID-19(62,63), additional sensitivity analyses did not show any significant Mendelian randomization between the general genetics of schizophrenia(64) and severe COVID-19 (see Supplementary Table 6). Activation of the classical, lectin and alternative complement pathways is associated with severity of COVID-19,(65–67) and microvascular injury and thrombosis in fatal cases.(67,68) Small non-randomised studies(69,70) have suggest the C5 inhibitor, eculizumab is a potential therapeutic for COVID-19. Finally in the MHC region, we also found evidence that HLA-DQA2 is causally associated with severe COVID-19 (OR = 2.29, 95% CI 1.94-2.64). In contrast to other HLA class II molecules, HLA-DQA2 is less polymorphic with a more discrete expression profile.(71)^,(72)^.

Turning to non-MHC proteins, we find proteinase 3 (PRTN3; OR = 2.48, 95% CI: 2.04-2.91) is causally associated with severe COVID-19. PRTN3 is a serine proteinase expressed in neutrophil granulocytes and a constituent of azurophil granules, that contain proteases and antibacterial proteins. PRTN3 is involved in neutrophil degranulation, highly overexpressed (29-fold) in naso-oropharyngeal samples of COVID-19 patients,(73) and associated with greater COVID-19 severity.(74) Neutrophil degranulation itself is dysregulated in COVID-19(75) while the myelopoiesis produces immature and dysfunctional neutrophils in severe COVID-19.(76) Given that PRTN3 is also implicated in immune dysregulation, sepsis and acute kidney injury,(77) this molecule may represent an interesting target to modulate immune responses against SARS-CoV-2.

The LDL receptor-related protein associated protein 1 (LRPAP1; OR = 1.73, 95% CI: 1.49-1.97) is involved as an escort protein in trafficking members of the LDL receptor family through the endoplasmic reticulum and Golgi apparatus preventing premature binding of ligands with these receptors.(78) In COVID-19, LDL cholesterol below or equal to 69 mg/dl has been associated with poor clinical outcomes.(79) Regarding a potential involvement in the development of neurological and cardiovascular symptoms, genomic variants in LRPAP1 have been associated with late-onset Alzheimer’s and Parkinson’s disease while serum anti-LRPAP1 is commonly used as a biomarker for atherosclerotic diseases.(80–82)

Our study has several limitations. Firstly, although we required confirmation of our findings by several Mendelian randomization methods, the *p*-value threshold for selecting genetic variants as instruments for our analyses was set at suggestive significance (*p* < 5 × 10^−6^) to allow enough to be identified for each protein, meaning that some of the genetic variants may not have true associations with protein levels. Secondly, some blood marker GWASs were excluded from our analyses due to unavailability, therefore, we may have missed associations with these markers. Thirdly, the severe COVID-19 GWAS used is of small sample size, reducing power to detect associations. However, it was a GWAS of the extreme phenotype of COVID-19 mortality or ventilator support, which may compensate for this.(83) Finally, our findings are limited to ancestral European populations due to data availability. Future endeavours should include participants from diverse ancestry groups.

Our results highlight the utility of applying large scale Mendelian randomization analyses to identify blood markers that may be causal for severe COVID-19. We demonstrate that TNNI3, PRTN3, HLADQA2, C4a-C4b and LRPAP1 may increase risk for severe COVID-19 while MHC1A and NCR3 may protect against severe COVID-19. These blood markers may play a role in the severity of COVID-19, indicating possible avenues to develop prognostic biomarkers and therapeutics for COVID-19.

## Data Availability

All data and software is publicly available via original sources.

## Acknowledgements

This study is supported by the National Institute for Health Research (NIHR) Mental Health Translational Research Collaboration (MH-TRC). The views expressed are those of the authors and not necessarily those of the NIHR or the Department of Health and Social Care.

## Author contributions

ABP, VM and CH collected, processed and analysed the data. ABP, VM, CH and GB wrote the manuscript. DM, LT and BM helped write the manuscript and added expert knowledge. This work was carried out as part of the COVID Clinical Neuroscience Study consortium.

## Data availability

All data and software is publicly available via original sources.

## Abbreviations

C4A: complement C4A (Rodgers blood group)
C4B: complement C4B (Chido blood group)
CREB3L4: cAMP responsive element binding protein 3 like 4
COVID-19: coronavirus disease 2019
DEFB119: beta-defensin 119
GWAS: genome-wide association study
HLA-DQA2: major histocompatibility complex (MHC), class II, DQ alpha 2
LRPAP1: low-density lipoprotein (LDL) receptor-related protein associated protein 1
MICA: MHC class I polypeptide-related sequence A
NCR3: natural cytotoxicity triggering receptor 3
OR: odds ratio
PRTN3: proteinase 3
TNNI3: troponin I3, cardiac type
SARS-CoV-2: severe acute respiratory syndrome coronavirus 2
SE: standard error
SNP: single nucleotide polymorphism

