## Supplementary Materials for "Proteome-wide Mendelian randomization identifies causal links between blood proteins and severe COVID-19"

Supplementary Table 1. A breakdown of all the studies from which inflammatory marker genome-wide association study (GWAS) data originated	2

Supplementary Table 2. Results from sensitivity analyses for all markers	6

Supplementary Table 3. A matrix of top SNPs from overlapping markers on chromosome 6	10

Supplementary Table 4. Details on the tissue, function and Covid-19 relevance of each significant blood biomarker	12

Supplementary Figure 1. A bubble plot representing genetically associated diseases and phenotypes with troponin I3, cardiac type	23

Supplementary Figure 2. A bubble plot representing disease and phenotype drug targets associated with TNNI3	25

Supplementary Table 5. Targets that are similar to TNNI3 (A) based on their disease association profiles	27

Supplementary Table 6. Mendelian randomization results between schizophrenia and severe COVID-19	28

### Supplementary Table 1. A breakdown of all the studies from which inflammatory marker genome-wide association study (GWAS) data originated

| **First author** | **Date** | **PMID** | **Link** | **Ancestry** | **Study name** | **Total N** | **Age** | **Number of males** | **Number of females** | **Number of markers** | **Platform** | **Normalization** | **Adjustment** |
| --- | --- | --- | --- | --- | --- | --- | --- | --- | --- | --- | --- | --- | --- |
| Wood | 2013 | 23696881 | <https://pubmed.ncbi.nlm.nih.gov/23696881/> | European (Italy) | InCHIANTI | 1210 | Mean = 68.2, Range = 21-102 | 540 | 670 | 30 proteins selected (out of 93) | N/A | inverse-normalized 2 times | age, sex |
| Suhre | 2017 | 28240269 | <https://pubmed.ncbi.nlm.nih.gov/28240269/> | European (southern Germany) | KORA F4 | 997 | N/A | 483 | 514 | 1124 | SOMAscan | inverse-normalized probe levels | age, sex, BMI |
| Ahola-Olli | 2017 | 27989323 | <https://pubmed.ncbi.nlm.nih.gov/27989323/> | European (Finland) | Cardiovascular Risk in Young Finns Study & FINRISK | 840-8293 | N/A | N/A | N/A | 41 | Bio-Rad’s premixed Bio-Plex Pro Human Cytokine 27-plex Assay and 21-plex Assay, and Bio-Plex 200 reader | normalized with inverse transformation. | age, sex, BMI, 10 first PC |
| Folkersen | 2017 | 28369058 | <http://dx.plos.org/10.1371/journal.pgen.1006706> | European (Finland, Sweden, France, Italy, Netherlands) | IMPROVE | 3394 | Mean = 64.5 (1) Range = 55-79 | N/A | N/A | 84 | OLINK | logarithmic | age, sex, recruitment center, protein analysis batch, smoking, diabetes and hypertension at baseline. |
| Sun | 2018 | 29875488 | <https://pubmed.ncbi.nlm.nih.gov/29875488/> | European (England) | INTERVAL | 3301 | Mean = 43.7 | 1686 | 1615 | 3622 | SOMAscan | logarithmic | age, sex, duration between blood draw and processing, 3 first PC |
| Sliz | 2019 | 31217265 | <https://www.ncbi.nlm.nih.gov/pubmed/?term=31217265> | European (Finland) | Northern Finland Birth Cohort, meta-analysis for 10 markers (same as Ahola) | 5284 - 13577 | 31 (2) | 2543 (2) | 2741 (2) | 16 (2) | Bio-Rad’s premixed Bio-Plex Pro Human Cytokine 27-plex Assay and 21-plex Assay, and Bio-Plex 200 reader | rank-based inverse transformation | age, sex, BMI, 10 first PC |
| Bretherik | 2020 | 32628676 | <https://www.ncbi.nlm.nih.gov/pmc/articles/PMC7337286/> | European (Scottland, Croatia) | ORCADES and CROATIA-Vis | 971-993 (ORCADES), 887-899 (Croatia-VIS) | Range 16-100 (ORCADES), 18-93 (Croatia-VIS) | N/A | N/A | 249 | OLINK | rank-based, inverse-normal transformed | age, sex, genotyping array (ORCADES), proteomics plate, plate row, column, length of sample storage, sease (ORCADES), 10 first PC |
| Scallop | 2020 | 33067605 | <https://www.nature.com/articles/s42255-020-00287-2#Abs1> | European (except STABILITY) | IMPROVE, STANLEY, EpiHealth, PIVUS, ULSAM, INTERVAL, LifeLines-DEEP, NSPHS, STABILITY, Estonian BB, ORCADES, VIS, MPP-RES | up to 21,758 (mean 17747) | N/A | N/A | N/A | 90 | OLINK | rank-based, inverse-normal transformed | depending on cohort |

(1) according to Strawbridge et al., (2014) ^84^

(2) for the Northern Finish Birth Cohort

PMID = PubMed ID

BMI = body mass index

EDTA = Ethylenediaminetetraacetic acid

PC = principal component

### Supplementary Table 2. Results from sensitivity analyses for all markers

| **Exposure** | **Outcome** | **Method** | **Number of SNPs** | **Beta** | **Standard Error** | **P-value** |
| --- | --- | --- | --- | --- | --- | --- |
| MICA | Severe_Covid | Robust adjusted profile score (RAPS) | 12 | -0.5637657 | 0.1596063 | 0.0004121 |
| MICA | Severe_Covid | Weighted median | 12 | -0.5072736 | 0.2029 | 0.0124153 |
| MICA | Severe_Covid | Inverse variance weighted | 12 | -0.5499291 | 0.149058 | 0.0002248 |
| MICA | Severe_Covid | MR Egger | 12 | -0.4572872 | 0.5187686 | 0.3987466 |
| HLA.DQA2 | Severe_Covid | Robust adjusted profile score (RAPS) | 14 | 1.0171603 | 0.289718 | 0.0004467 |
| HLA.DQA2 | Severe_Covid | Weighted median | 14 | 1.1714867 | 0.3626369 | 0.0012359 |
| HLA.DQA2 | Severe_Covid | Inverse variance weighted | 14 | 0.9594586 | 0.2651303 | 0.000296 |
| HLA.DQA2 | Severe_Covid | MR Egger | 14 | 1.495111 | 0.5552437 | 0.0195715 |
| LRPAP1 | Severe_Covid | Robust adjusted profile score (RAPS) | 18 | 0.5503255 | 0.221648 | 0.0130325 |
| LRPAP1 | Severe_Covid | Weighted median | 18 | 0.6909397 | 0.2688378 | 0.0101669 |
| LRPAP1 | Severe_Covid | Inverse variance weighted | 18 | 0.536829 | 0.2096652 | 0.0104549 |
| LRPAP1 | Severe_Covid | MR Egger | 18 | 0.6107214 | 0.3543689 | 0.1040813 |
| C4A.C4B | Severe_Covid | Robust adjusted profile score (RAPS) | 25 | 0.8685084 | 0.2210557 | 0.0000853 |
| C4A.C4B | Severe_Covid | Weighted median | 25 | 0.8404732 | 0.2886189 | 0.0035906 |
| C4A.C4B | Severe_Covid | Inverse variance weighted | 25 | 0.7667997 | 0.2253834 | 0.0006685 |
| C4A.C4B | Severe_Covid | MR Egger | 25 | 0.7054519 | 0.4650157 | 0.1428797 |
| CREB3L4 | Severe_Covid | Robust adjusted profile score (RAPS) | 10 | 0.4709533 | 0.221723 | 0.033665 |
| CREB3L4 | Severe_Covid | Weighted median | 10 | 0.4552106 | 0.239626 | 0.0574763 |
| CREB3L4 | Severe_Covid | Inverse variance weighted | 10 | 0.4684829 | 0.2112441 | 0.0265731 |
| CREB3L4 | Severe_Covid | MR Egger | 10 | 0.4982686 | 0.3027253 | 0.1383932 |
| PRTN3 | Severe_Covid | Robust adjusted profile score (RAPS) | 14 | 1.0154735 | 0.2761558 | 0.0002358 |
| PRTN3 | Severe_Covid | Weighted median | 14 | 1.1943379 | 0.3190539 | 0.0001816 |
| PRTN3 | Severe_Covid | Inverse variance weighted | 14 | 0.9224542 | 0.2596129 | 0.0003806 |
| PRTN3 | Severe_Covid | MR Egger | 14 | 1.1079149 | 0.5141108 | 0.0521742 |
| TNNI3 | Severe_Covid | Robust adjusted profile score (RAPS) | 7 | 1.768093 | 0.8058205 | 0.0282244 |
| TNNI3 | Severe_Covid | Weighted median | 7 | 1.509658 | 0.6988436 | 0.0307554 |
| TNNI3 | Severe_Covid | Inverse variance weighted | 7 | 1.729311 | 0.5097016 | 0.0006918 |
| TNNI3 | Severe_Covid | MR Egger | 7 | 1.1032 | 1.6119381 | 0.5241529 |
| DEFB119 | Severe_Covid | Robust adjusted profile score (RAPS) | 14 | 0.4214074 | 0.3714694 | 0.2566127 |
| DEFB119 | Severe_Covid | Weighted median | 14 | 0.2370707 | 0.3612275 | 0.5116364 |
| DEFB119 | Severe_Covid | Inverse variance weighted | 14 | 0.4002511 | 0.349509 | 0.2521341 |
| DEFB119 | Severe_Covid | MR Egger | 14 | 0.2904774 | 0.7046881 | 0.687456 |
| NCR3 | Severe_Covid | Robust adjusted profile score (RAPS) | 12 | -1.228803 | 0.3435943 | 0.0003485 |
| NCR3 | Severe_Covid | Weighted median | 12 | -1.455385 | 0.4670759 | 0.0018335 |
| NCR3 | Severe_Covid | Inverse variance weighted | 12 | -1.201865 | 0.3158843 | 0.0001419 |
| NCR3 | Severe_Covid | MR Egger | 12 | -2.065071 | 0.7680217 | 0.0227458 |

Number of SNPS = nsnp / Beta = b / Standard Error = se / P-value = pval

C4A = complement C4A (Rodgers blood group), C4B = complement C4B (Chido blood group), CREB3L4 = cAMP responsive element binding protein 3 like 4, DEFB119 = beta-defensin 119, HLA-DQA2 = major histocompatibility complex (MHC), class II, DQ alpha 2, LRPAP1 = low-density lipoprotein (LDL) receptor-related protein associated protein 1, MICA = MHC class I polypeptide-related sequence A, NCR3 = natural cytotoxicity triggering receptor 3, PRTN3 = proteinase 3, TNNI3 = troponin I3, cardiac type.

### Supplementary Table 3. A matrix of top SNPs from overlapping markers on chromosome 6

| RS_number | rs79959614 | rs73391152 | rs12190662 | rs17188113 | rs6926224 | rs12662617 | rs11757571 | rs143439747 | rs16869834 | rs9271547 | rs3134971 | rs28724893 | rs3097665 |
| --- | --- | --- | --- | --- | --- | --- | --- | --- | --- | --- | --- | --- | --- |
| rs79959614 | 1 | 0.453 | 0.003 | 0.004 | 0.002 | 0.002 | 0.001 | 0.001 | 0 | 0.012 | 0.002 | 0.008 | 0.002 |
| rs73391152 | 0.453 | 1 | 0.006 | 0.012 | 0.002 | 0.001 | 0.002 | 0 | 0.002 | 0.009 | 0.003 | 0.013 | 0 |
| rs12190662 | 0.003 | 0.006 | 1 | 0.115 | 0.015 | 0.003 | 0.001 | 0.001 | 0.001 | 0.009 | 0.003 | 0.014 | 0.001 |
| rs17188113 | 0.004 | 0.012 | 0.115 | 1 | 0.017 | 0.001 | 0.003 | 0.001 | 0.014 | 0.012 | 0.014 | 0.007 | 0.008 |
| rs6926224 | 0.002 | 0.002 | 0.015 | 0.017 | 1 | 0.024 | 0.002 | 0.049 | 0.002 | 0.011 | 0.001 | 0.009 | 0.001 |
| rs12662617 | 0.002 | 0.001 | 0.003 | 0.001 | 0.024 | 1 | 0.006 | 0.005 | 0.003 | 0.001 | 0.019 | 0.015 | 0.004 |
| rs11757571 | 0.001 | 0.002 | 0.001 | 0.003 | 0.002 | 0.006 | 1 | 0.003 | 0.003 | 0.01 | 0.013 | 0 | 0.001 |
| rs143439747 | 0.001 | 0 | 0.001 | 0.001 | 0.049 | 0.005 | 0.003 | 1 | 0.047 | 0.005 | 0.006 | 0.008 | 0.001 |
| rs16869834 | 0 | 0.002 | 0.001 | 0.014 | 0.002 | 0.003 | 0.003 | 0.047 | 1 | 0.008 | 0.003 | 0.001 | 0.001 |
| rs9271547 | 0.012 | 0.009 | 0.009 | 0.012 | 0.011 | 0.001 | 0.01 | 0.005 | 0.008 | 1 | 0.127 | 0.015 | 0.012 |
| rs3134971 | 0.002 | 0.003 | 0.003 | 0.014 | 0.001 | 0.019 | 0.013 | 0.006 | 0.003 | 0.127 | 1 | 0.102 | 0 |
| rs28724893 | 0.008 | 0.013 | 0.014 | 0.007 | 0.009 | 0.015 | 0 | 0.008 | 0.001 | 0.015 | 0.102 | 1 | 0.001 |
| rs3097665 | 0.002 | 0 | 0.001 | 0.008 | 0.001 | 0.004 | 0.001 | 0.001 | 0.001 | 0.012 | 0 | 0.001 | 1 |

This table displays the top SNPs used as instruments in blood proteins associated with higher/lower odds of severe COVID-19, which are located in close proximity on chromosome 6. These SNPs are primarily responsible for the observed blood protein - COVID-19 association. SNPs are ordered according to chromosome position. Color codes are displayed below. HLA-DQA2 = major histocompatibility complex (MHC), class II, DQ alpha 2, C4A = complement C4A (Rodgers blood group), C4B = complement C4B (Chido blood group), MICA = MHC class I polypeptide-related sequence A, NCR3 = natural cytotoxicity triggering receptor 3.

| HLA-DQA2 | C4a-C4b |
| --- | --- |
| MICA | NCR3 |

### Supplementary Table 4. Details on the tissue, function and Covid-19 relevance of each significant blood biomarker

| **Name** | **Gene** | **Tissue** | **Function** | **Covid-19 hypothesis** |
| --- | --- | --- | --- | --- |
| Troponin I3, cardiac type | TNNI3 | Heart muscle tissue (only) | One of three proteins that is part of the cardiac troponin protein complex  Associated with the sarcomere  Part of the muscle contraction process  Marker for myocardial infarctions | Cardiomyopathie, either hypertrophic or dilatative  May increase risk for sudden heart failure  Meta-analysis as biomarker and COVID-19 → increased in severe forms ^85^  Myocardial damage ^86^  Troponin I is an independent predictor of mortality in COVID-19 ^44^  Review for testing ^87^ |
| Proteinase 3 | PRTN3 | Neutrophil granulocytes | Serine protease enzyme  Degrades elastin, fibronectin, laminin, vitronectin, and collagen types I, III, and IV (in vitro)  Exact function is unknown  By cleaving and activating receptor F2RL1/PAR-2, enhances endothelial cell barrier function and thus vascular integrity during neutrophil transendothelial migration  May play a role in neutrophil transendothelial migration, probably when associated with CD177  Proteolytic generation of antimicrobial peptides  Target for anti-neutrophil cytoplasmic antibodies (ANCAs) of the c-ANCA (cytoplasmic subtype)  Associated with Wegener Granulomatosis | Endothelial migration of neutrophils → vasculitis association  Increased expression of PRTN3 and neutrophil activation in COVID-19 ^76^  Increased neutrophil count in severe COVID-19 ^77^  Altered in nasopharyngal swabs of COVID-19 patients ^74^ |
| Major Histocompatibility Complex, Class II, DQ Alpha 2 | HLA-DQA2 | Antigen presenting cells (B lymphocytes, dendritic cells, macrophages) | Protein forms a heterodimer with a class II beta chain  Located in intracellular vesicles  Present antigenic peptides on the cell surface to be recognized by CD4 T-cells (speculative)  Peptides presented by MHC class II molecules are generated mostly by degradation of proteins that access the endocytic route → exogenous  Associated with podoconiosis & rheumatoid arthritis | Autoantigen presentation may lead to cytokine storm (speculative)  Gene associated with forced expiratory volume (FEV1) in previous GWAS (lung function) ^88^  Also identified in MAGMA analysis ^89^  HLA-DQA1 is downregulated (genes adjacent) ^90^ |
| Beta-defensin 119 | DEFB119 | Testis and epididymis | Member of the beta subfamily of defensins  Antimicrobial peptides  Protective against infections  Associated with spastic paraplegia 20 |  |
| cAMP Responsive Element Binding Protein 3 Like 4 | CREB3L4 | Omnipresent | Protein with a transmembrane domain  Endoplasmic reticulum membrane  Transcriptional activator  Contains a dimerization domain  A number of processing pathways including protein processing  Different isoforms  Unfolded protein response (UPR)  Interacts with androgen receptor  Associated with prostate and breast cancer |  |
| Complement C4A (Rodgers Blood Group) | C4A | Alveolar cells type 2, club cells, alveolar cells type 2, ciliated cells; enhanced in liver | Acidic form of complement factor 4  Single chain precursor  Proteolytically cleaved into a trimer of alpha, beta, and gamma chains prior to secretion  Trimer provides a surface for interaction between the antigen-antibody complex and other complement components  Alpha chain is cleaved to release C4 anaphylatoxin, an antimicrobial peptide and a mediator of local inflammation  It induces the contraction of smooth muscle, increases vascular permeability and causes histamine release from mast cells and basophilic leukocytes  Deficiency of this protein is associated with systemic lupus erythematosus and type I diabetes mellitus  Localizes to the major histocompatibility complex (MHC) class III region on chromosome 6  Varying haplotypes of this gene cluster exist, such that individuals may have 1, 2, or 3 copies of this gene  Different isoforms have been found for this gene  Non-enzymatic component of C3 and C5 convertases and thus essential for the propagation of the classical complement pathway  Covalently binds to immunoglobulins and immune complexes and enhances the solubilization of immune aggregates and the clearance of IC through CR1 on erythrocytes  C4A isotype is responsible for effective binding to form amide bonds with immune aggregates or protein antigens | C3 and C4 is decreased in severe COVID-19 patients ^91^ |
| Complement C4B (Chido Blood Group) | C4B |  | See above  C4B isotype catalyzes the transacylation of the thioester carbonyl group to form ester bonds with carbohydrate antigens |  |
| LDL Receptor Related Protein Associated Protein 1 | LRPAP1 | Widely expressed | Chaperone protein  Glycoprotein  Trafficking of LDL receptors, including LRP1 and LRP2  Facilitates its proper folding and localization  Binds to alpha-2-macroglobulin receptor  Inhibits binding  May prevent receptor aggregation and degradation in the endoplasmic reticulum  Associated with myopia 23 | LDL is associated with poor clinical outcomes in COVID-19 ^92^  MR study: LDL → Covid-19 ^93^ |
| MHC Class I Polypeptide-related Sequence A | MICA | Lung, endometrium and 25 other tissues  No central nervous system expression | Highly polymorphic major histocompatibility complex class I chain-related protein A  Expressed on the cell surface  Glycoprotein  Although unlike canonical class I molecules it does not seem to associate with beta-2-microglobulin  Does not bind peptides  Ligand for the NKG2-D type II integral membrane protein receptor/killer cell lectin like receptor K1  Binding to the KLRK1/NKG2D receptor leads to cell lysis  Functions as a stress-induced antigen that is broadly recognized by intestinal epithelial gamma delta T cells  Triggered by malignant transformation such as DNA damage and accumulation of misfolded proteins  Also recognized by nature killer cells and CD8+ alpha beta T cells  Effector cytolytic responses of T cells  Associated with psoriasis 1 and psoriatic arthritis, spondylitis, and multiple myeloma  Alternative splicing of this gene results in multiple transcript variants  No ortholog in mice | Expression is affected by infectious agents, such as the human cytomegalovirus, human adenovirus, M. tuberculosis, E. coli, or human papillomavirus (HPV)  Association with diarrhea |
| Natural Cytotoxicity Triggering Receptor 3 | NCR3 | Selectively expressed by all resting and activated NK cells and weakly expressed in spleen | Cell membrane receptor of natural killer/NK cells that is activated by binding of extracellular ligands including BAG6 and NCR3LG1  Stimulates NK cells cytotoxicity toward neighboring cells producing these ligands.  May aid NK cells in the lysis of tumor cells  Interacts with CD3-zeta (CD247), a T-cell receptor signalling component  A single nucleotide polymorphism in the 5' untranslated region of this gene has been associated with mild malaria susceptibility  Three transcript variants encoding different isoforms have been found for this gene. |  |

C4A = complement C4A (Rodgers blood group), C4B = complement C4B (Chido blood group), CREB3L4 = cAMP responsive element binding protein 3 like 4, DEFB119 = beta-defensin 119, HLA-DQA2 = major histocompatibility complex (MHC), class II, DQ alpha 2, LRPAP1 = low-density lipoprotein (LDL) receptor-related protein associated protein 1, MICA = MHC class I polypeptide-related sequence A, NCR3 = natural cytotoxicity triggering receptor 3, PRTN3 = proteinase 3, TNNI3 = troponin I3, cardiac type.

### Supplementary Figure 1. A bubble plot representing genetically associated diseases and phenotypes with troponin I3, cardiac type

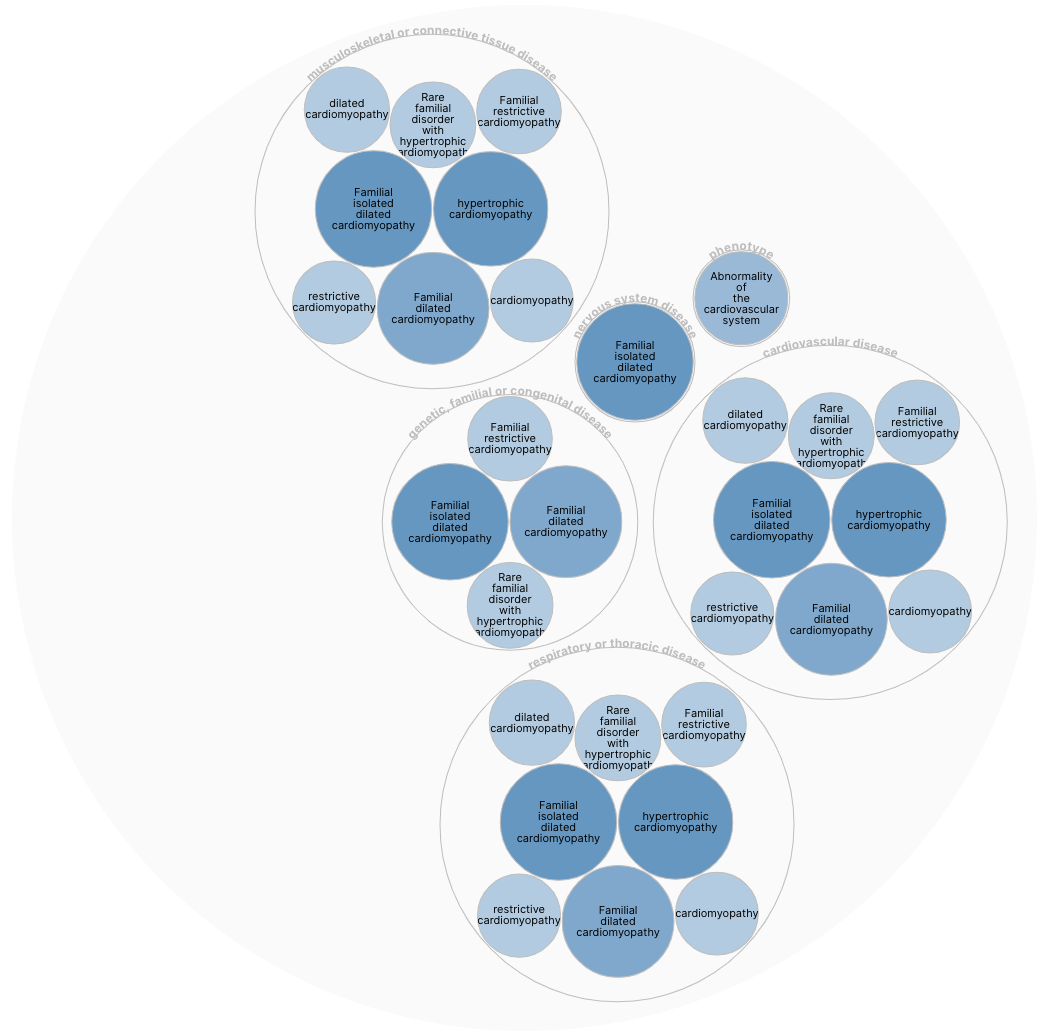

This bubble plot represents the genetically associated diseases and phenotypes with troponin I3, cardiac type (TNNI3). Data was obtained using the Open Targets Platform (<https://www.targetvalidation.org/>) and the minimum association score was set at the predetermined threshold of 0.1

### Supplementary Figure 2. A bubble plot representing disease and phenotype drug targets associated with TNNI3

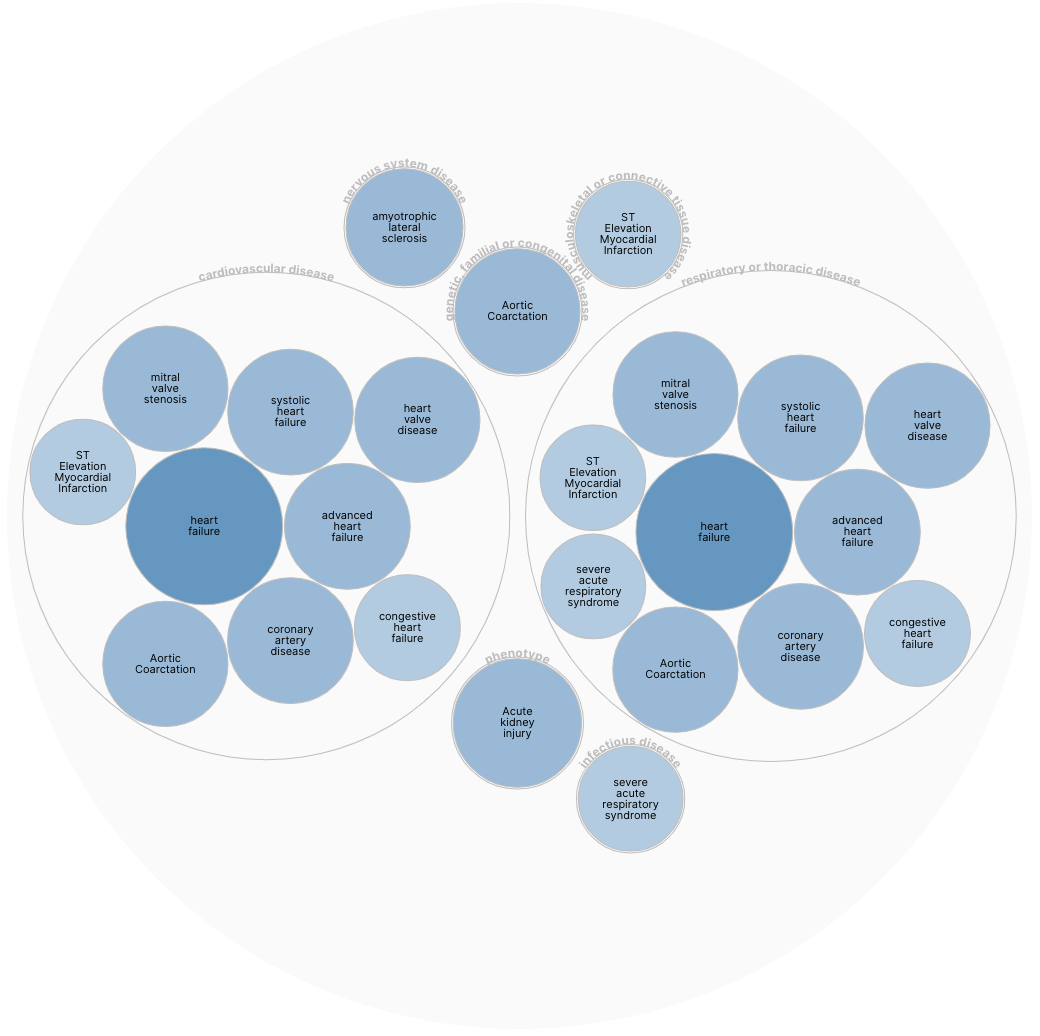

This bubble plot represents the diseases and phenotype drug targets associated with troponin I3, cardiac type (TNNI3). Data was obtained using the Open Targets Platform (<https://www.targetvalidation.org/>) and the minimum association score was set at the predetermined threshold of 0.1. The associated drug in all instances is levosimendan.

### Supplementary Table 5. Targets that are similar to TNNI3 (A) based on their disease association profiles

| **Related Target (B)** | **Similarity Score** | **A not B** | **A and B** | **B not A** |
| --- | --- | --- | --- | --- |
| MB | 0.53100492 | 37 | 24 | 21 |
| NPPA | 0.51588324 | 31 | 30 | 47 |
| ACE2 | 0.4983143 | 32 | 29 | 52 |
| TRIM63 | 0.4978442 | 42 | 19 | 13 |

TNNI3 = troponin I3, cardiac type; MB = myoglobin; NPPA = natriuretic peptide A; ACE2 = angiotensin I converting enzyme 2; TRIM63 = tripartite motif containing 63. “A” denotes TNNI3 and “B” denotes the target protein. “A not B” describes diseases associated with TNNI3 but not the related target; “A and B” describes shared disease associations; “B not A” describes diseases associated with the related target but not TNNI3. The similarity score is computed via the Open Targets Platform ([https://www.targetvalidation.org](https://www.targetvalidation.org/)) and encompasses the relative occurrence of a target-disease evidence, the magnitude or strength of the effect described by the evidence, and the overall confidence for the observation that generates the target-disease evidence. Other columns indicate how many diseases are associated with the proteins in question in that category.

### Supplementary Table 6. Mendelian randomization results between schizophrenia and severe COVID-19

| **Exposure** | **Outcome** | **Beta** | **SE** | **P** | **SNPs** | **P-value threshold** |
| --- | --- | --- | --- | --- | --- | --- |
| Schizophrenia | Severe COVID-19 | 0.017444 | 0.119813 | 0.884242 | 276 | 5.00E-08 |
| Severe COVID-19 | Schizophrenia | -0.0039291 | 0.00314603 | 0.211698 | 10 | 5.00E-06 |

Note: SE = standard error, SNPs = number of single nucleotide polymorphisms in common between the exposure and outcome, used for the Mendelian randomization analysis; OR = odds ratio. The P-value threshold signifies the threshold below which SNPs were obtained for the analysis.
